# Food Insecurity in the Households of Children with Autism Spectrum Disorders and Intellectual Disabilities in the U.S.: Analysis of the National Survey of Children’s Health Data 2016 - 2018

**DOI:** 10.1101/2021.03.29.21254546

**Authors:** Arun Karpur, Vijay Vasudevan, Angela Lello, Thomas W. Frazier, Andy Shih

**Affiliations:** Autism Speaks, Inc., 1060 State Road, Princeton, NJ 08540

**Keywords:** Autism Spectrum Disorders (ASD), Food Insecurity, COVID19, Sociotype

## Abstract

Individuals with Autism Spectrum Disorder and co-occurring Intellectual Disabilities (ASD + ID) experience substantial challenges in accessing needed supports. This research aimed to understand the prevalence and factors associated with food insecurity among families of children with ASD + ID. Utilizing the National Survey of Children’s Health (2016-18) data, this paper illustrated that the households of children with ASD + ID were about two times more likely to be food insecure than the households of children without disabilities. Further, the households of children with ASD were 1.5 times more likely, and those with other disabilities were 1.3 times more likely to be food insecure than the households of children without disabilities. Implications of these findings in the context of the COVID19 pandemic are discussed.

**Lay Abstract:** Families of children with ASD are more likely to experience financial strain and resulting food insecurity due to additional cost of care, disparate access to needed services, and loss of income resulting from job loss. Utilizing nationally representative data, this analysis indicates that the families of children with ASD + ID are twice as likely to experience food insecurity than families of children without disabilities after adjusting for various factors. Several factors, ranging from state-level policies such as Medicaid expansion to individual-level factors such as higher utilization of emergency room services, were associated with the higher prevalence of food insecurity in families of children with ASD + ID. Implications of these findings on programs and policies supporting families in the COVID19 pandemic are discussed.

---

More than 54 million Americans are estimated to be food insecure in the COVID19 pandemic (Hake et al., 2020). Food insecurity is a household level construct defined as the inability or uncertainty in accessing adequate food for an active and healthy lifestyle. Since the Great Recession of 2008-2009, annually, 12% - 15% (or 37 million) individuals in the U.S. have experienced food insecurity (Hake et al., 2020). However, certain groups of households experience higher levels of food insecurity: i.e., those with children (13.6%), Black, non-Hispanic and Hispanic households (19.1% and 15.6% respectively), and households with a working-age adult with a disability (38%) (Coleman-Jansen, 2020; Coleman-Jensen & Nord, 2013; Coleman-Jensen, Rabbitt, Gregory, & Singh, 2020; Schwartz, Buliung, & Wilson, 2019).

Research on the prevalence and factors associated with food insecurity in households with children with autism spectrum disorder (ASD) is very limited. As financial and social vulnerabilities impacting families of individuals with ASD are, on average, higher compared to those with other disabilities and those without disabilities (Rogge & Janssen, 2019), it is likely that they also experience higher levels of food insecurity.

## Vulnerabilities among Individuals with ASD

ASD is a life-long neurodevelopmental condition characterized by social and communication disorders (Maenner, Shaw, Baio, & al., 2020). Individuals with ASD also experience co-occurring conditions such as intellectual disabilities, epilepsy, mental health conditions, gastrointestinal disorders, sleep disorders, etc., requiring access to specialist and other forms of high-cost services (Soke, Maenner, Christensen, Kurzius-Spencer, & Schieve, 2018). A greater proportion of families of children with ASD indicate higher unmet healthcare needs compared to other children with special healthcare needs and those without any disabilities irrespective of their level of access to health insurance and medical care (A. Karpur, Lello, Frazier, Dixon, & Shih, 2018; Soke et al., 2018). From a life course perspective, individuals with ASD are less likely to complete high school, engage in work, and experience higher instances of engagement with law enforcement than their peers without disabilities (Roux, Rast, Garfield, Anderson, & Shattuck, 2020). The systemic barriers in access to needed services, and lack of opportunities to engage in community-based activities are also likely to contributed to a substantially lower life expectancy among individuals with ASD (Guan & Li, 2017; Hirvikoski et al., 2016).

The total lifetime cost of care for individuals with ASD is estimated to range from $2.4 million to $3.2 million, and it is forecasted that by 2025 the cost of health care alone will be $461 billion (Lavelle et al., 2014). Families of individuals with ASD are estimated to spend $4,000 more in out-of-pocket health care expenses annually than those without disabilities (Rogge & Janssen, 2019). The families also experience economic loss of up to $20,000 annually from lost productivity and parents of children with ASD are 2.5 times more likely to stop working or reduce work hours in order to care for their child (A Karpur, 2019; McCall & Starr, 2016; Rogge & Janssen, 2019). Families experience high levels of social stigma and isolation contributing to their mental health stress (Frazier, Hyland, Markowitz, Speer, & Diekroger, 2020; Markowitz et al., 2016; Werner & Shulman, 2014). Further, children with ASD have a higher probability of exposure to adverse childhood experience or ACEs (e.g., witnessing domestic violence, parents with history of incarceration, experiencing discrimination, etc.) which predisposes them to additional health risks including food insecurity (Chilton, Knowles, Rabinowich, & Arnold, 2015; Jackson, Chilton, Johnson, & Vaughn, 2019; Sun et al., 2016; Testa & Jackson, 2020). Increasing identification of ASD in low-income and minority populations likely contributes to an overall higher proportion of social and economic vulnerability among these families (Nevison & Parker, 2020).

Macro-level factors such as living in service-rich environment or in states that expanded Medicaid eligibility under the Affordable Care Act or having access to Medicaid Home and Community Based (HCBS) waiver services improves overall well being and food insecurity among individuals with ASD. For example, the probability of food insecurity reduced by over 13% in the states that expanded Medicaid eligibility (Gracie Himmelstein, 2019). Additionally, expansion of access to services under HCBS waivers has substantially reduced unmet healthcare needs in children with ASD and improved overall family functioning, especially among those who could not qualify for Medicaid (Leslie, Iskandarani, Dick, et al., 2017; Leslie, Iskandarani, Velott, et al., 2017).

### Sociotype as a framework for studying food insecurity

Food insecurity is a continuum and is not easily measured in objective terms using economic or calorie intake indicators (Webb et al., 2006). Radimer et al. (1992), using a qualitative approach to understanding the food insecurity among women in rural New York, illustrated that food insecurity was a “managed process,” where households responded idiosyncratically with some commonalities. When these households’ livelihoods were threatened, they experienced “worry” about obtaining sufficient food and deployed different strategies to improve their access to food. When these conditions continued, the households compromised on their quality of food, and children’s food quality and quantity were compromised only in extreme conditions (Radimer, Olson, Greene, Campbell, & Habicht, 1992). Any breakdown in the chain of availability to accessibility to utilization fosters food insecurity. Thus, some low-income households may be food insecure but not experience hunger, and others are likely to live in extreme duress. However, even the marginal form of food insecurity – i.e., a perception of uncertainty in accessing desired quantity and quality of food – has detrimental impact on the physical and mental well being of households (Cook et al., 2013).

The conceptual framing of sociotype helps to understand factors related to this continuum of food insecurity expressed as uncertainty in access and means for acquiring sufficient quality and quantity of safe and nutritious food (Berry, Bachar, Baras, & De Geest, 2017; Peng, Dernini, & Berry, 2018). Extending the Bronfenbrenner’s ecological model (Bronfenbrenner & Ceci, 1994; Vélez-Agosto, Soto-Crespo, Vizcarrondo-Oppenheimer, Vega-Molina, & García Coll, 2017), Engel’s biopsychosocial model (Adler, 2009), and the salutogenesis framework by Antonovsky (Vinje, Langeland, & Bull, 2017), the sociotype explains dynamic, reciprocal, and complex inputs from the environment that interact with the genotype (not studied in this analysis) leading to the phenotypic expression of food insecurity (Berry et al., 2017; Peng et al., 2018). By adding the intra-, and inter-personal inputs to broader ecological context, sociotype helps to uncover “deep social structures” (Chapais, 2011) describing the patterns of food insecurity in populations across contexts (Coates et al., 2006).

The sociotype construct consists of three domains: Individual Health (IH), Relationships (R), and Context (C). The IH domain consists of the physical, mental, and spiritual well-being of a person, including cumulative effects of any adverse experiences; the R domain consists of interpersonal relationships with family, parents, friends, professionals, etc.; and the C domain consists of socio-economic conditions, neighborhoods, social and political situations (see Fig. 1). Families experiencing food insecurity (or any form of stress) usually make their initial or primary assessment of the extent of stress and develop strategies to address them. The sociotype, drawing on family’s prior experiences, social networks, and access to resources, continually shapes their cognitive, emotional, and rational considerations leading to experiencing varying degrees of food insecurity (J. T. Denney, Kimbro, & Sharp, 2018; Mesoudi et al., 2017). While this analysis does not aim to empirically identify specific ‘sociotypes’ in the households of children with ASD, this research benefits from the organization of conceptual domains in the sociotype framework. Further, studying incremental contribution of variables across the IH, R, and C domains, the analysis aims to highlight their relevance in developing mitigating programs and policies to ensure food security among vulnerable families of children with ASD.

**. Figure 1.**
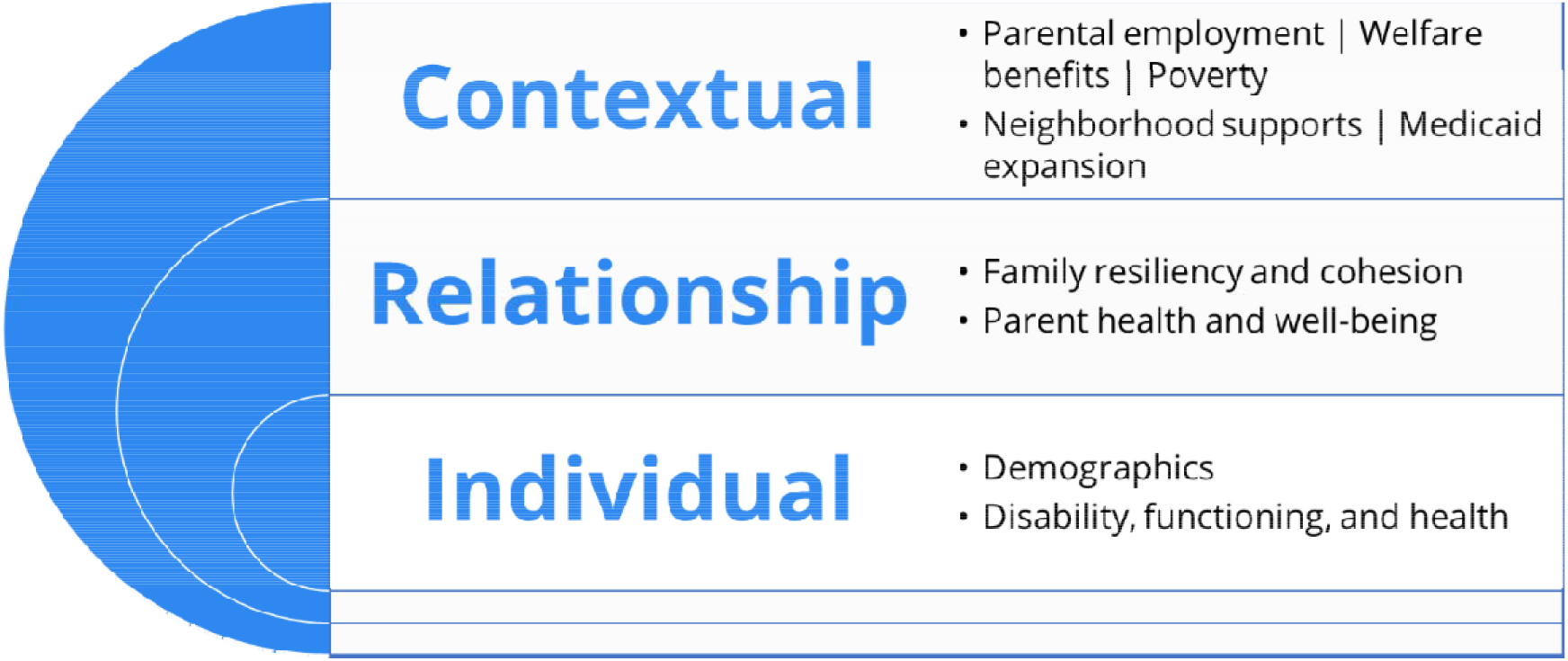
Variables across the sociotype domains explaining food insecurity.

The purpose of this research is to understand how individual health, relationships, and contextual factors impact the likelihood of being food insecure in families of individuals with ASD + ID and those with ASD only in comparison with those without disabilities utilizing nationally existing survey data.

## Methods

### Data source

Three years of data from the National Survey of Children’s Health (2016 – 2018) were combined for this analysis. Briefly, the NSCH is a nationally representative sample survey of children’s health administered annually by the U.S. Census Bureau for the Maternal and Child Health Bureau (MCBH), Health Resources and Services Administration (HRSA). This survey combines prior two separate surveys – the National Survey of Children’s Health (NSCH) and the National Survey of Children with Special Health Care Needs (NSCSHCN). The NSCH survey collects data from a nationally-representative households using: (1) a household screener questionnaire to capture demographics and special health care needs characteristics (aided with an oversampling of children with special health care needs); and (2) an age-based topical questionnaire (T1 for 0 to 5-year old, T2 for 6 to 11 year old, and T3 for 12 to 17 year old) that combines core elements from the NSCH and the NSCSHCN.

The overall response rates for the NSCH surveys in 2016 – 2018 range from about 41% - 43%. The survey weights are assigned to each respondent to obtain a population-level estimate. Importantly, the final survey weights are adjusted for nonrespondents to minimize the nonresponse bias in the analysis. Individual survey weights for each year were divided by the number of years of the survey data combined to derive nationally representative estimates. Further guidance on methodology and creating multi-year estimates can be found at (https://www2.census.gov/programs-surveys/nsch/technical-documentation/methodology/NSCH-Guide-to-Multi-Year-Estimates.pdf). No IRB approval was required as the analysis utilized publicly available datasets.

### Comparison groups

The children of parents responding affirmatively to the question, “Has a doctor or other health care provider EVER told you that this child has Autism or Autism Spectrum Disorder (ASD)? Include diagnoses of Asperger’s Disorder or Pervasive Developmental Disorder (PDD)” were classified as children with ASD. Further, the children of parents responding affirmatively to the question “Has a doctor or other health care provider EVER told you that this child has Intellectual Disability (formerly known as Mental Retardation)?” were classified as children with Intellectual Disabilities (ID). Additionally, the children classified as not having ASD but with a positive response to the CSHCN screener were classified as children with other disabilities, and finally, children without ASD and CSHCN were classified as children without disabilities. Four mutually exclusive groups were created to include children with ASD + ID (N = 447), children with ASD only (N = 2,238), children with other disabilities (N = 20,763), and children without disabilities (78,781).

### Dependent variable – food insecurity

Within the NSCH topical questionnaire, families were asked, “Which of these statements best describes the food situation in your household in the past 12 months?” Families responding to the statements “Sometimes we could not afford enough to eat” and “Often we could not afford to eat” were identified as food insecure. Additionally, families that also indicated that they “Somewhat often” or “Very often” found it hard to get by on family’s income to cover basic needs such as food or housing were considered to be food insecure. Based on this approach, 22% of responding households were food insecure.

### Independent variables

In addition to the disability groupings, several factors based on the sociotype framework were considered relevant for the analysis.

#### The individual health (IH) factors

included race/ethnicity, frequency of emergency room visits, being absent at school for 11 or more days in the past 12 months, and child’s exposure to adverse childhood experiences. Child’s exposure to adverse childhood experiences was self-reported by primary caregivers and was based on a set of items asking “to the best your knowledge, has (CHILD) EVER experienced any of the following?” and the list of adverse events such as parent divorce, parent history of incarceration or exposure to domestic violence, and experience of discrimination.

#### Relationship factors (R)

included the Family Resiliency and Connection Index (FRCI), access to emotional support, and perceived parental physical and mental health. Respondents that indicated “yes” when asked, “During the past 12 months was there someone that you could turn to for day-to-day emotional support with parenting or raising children?” were indicated as having access to emotional support. The FRCI was created based on six items indicating family resiliency (four items) and parent-child connection (two items) (Bethell, Gombojav, & Whitaker, 2019).

#### Context-level factors(C)

included parental employment status, gaps in child’s health insurance coverage, receipt of public assistance (including food or cash benefits), family’s estimated federal poverty level, living in a supportive neighborhood, and living in states that expanded Medicaid. A supportive neighborhood was identified based on the extent of agreement to items describing if the people “in the neighborhood help each other out,” “watch each other’s children,” and if they perceive that the child “is safe in the neighborhood,” and they know “where to go to get help in our community.” The family poverty level was created as a binary variable, where families below 200% federal poverty level were classified as economically vulnerable and those above 200% federal poverty level were classified as not economically vulnerable. This classification provides a meaningful way to group families, where those living under 200% FPL are more likely to be eligible for various welfare benefits and considered low-income families compared with those above 200 % FPL (Zedlewski, Chaudry, & Simms, 2009). Further, the state that expanded Medicaid access by 2016 under the ACA was identified as a binary variable [Medicaid expanded states = 1; 0 otherwise].

### Statistical analysis

The univariate analyses examined relationships between the factors across the IH, R, and C domains with food insecurity as well as their distribution across the comparison groups (i.e., children with ASD/ID, ASD, other disabilities, and those with no disabilities). Multiple logistic regression models, with food insecurity as a binary dependent variable, were constructed. First, the model examined the association between factors in the IH domain and food insecurity. Other groups of variables were added in subsequent models, and their extent of contributions in explaining variation in outcome variables was ascertained using the Tjur R-square (Tjur, 2009). The c-statistic for each of the models was also assessed for understanding model fit.

Briefly, the Tjur R-square statistic or the Coefficient of Determination (D) is calculated as follows. First, a logistic regression model is fit to the data and predicted probabilities for cases and non-cases are output. Then, histograms of the predicted probabilities are calculated for the cases and non-cases, and the mean value for each group of the histograms are computed. The difference in the mean value for cases (*π*_l_) and non-cases (*π*_0_) represents D and it is equivalent to the R-squared statistic (Kvalseth, 1985).

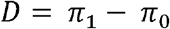

Additional proofs and methods are described in Tjur (2009). Examining the percent change in the R-squared value across the consecutive nested models demonstrates an incremental increase in the extent of variation predicted by the set of independent variables. The Tjur R-square has desirable properties compared to other R-square values in logistic regression, as it is not dependent on a particular estimation method.

### Community involvement

The lead author and co-authors work for a leading international organization advocating for the rights of people with ASD and their families. None of the authors are autistic, but the analysis was benefited through review from autistic advisory board members.

## Results

### Bivariate analysis

A substantially higher proportion of children with ASD + ID were food insecure (44%), followed by children with ASD only (40%), children with other disabilities (33%), and those with no disabilities (20%). Table 1 illustrates relationship of the independent variables associated with the likelihood of food insecurity. Children with ASD+ID were three times more likely to be food insecure compared with their peers without any disabilities (OR = 3.1; 95% CI: 1.7 – 5.3). Further children with ASD only were more than two times (OR = 2.3; 95% CI: 1.8 – 3.1), and those with other disabilities were 1.9 times more likely to be food insecure compared with their peers without disabilities (OR = 1.9; CI: 1.8 – 2.1). Similarly, children belonging to race/ethnic minorities, those missing school more than 11 days a year, those with a higher frequency of emergency room visits, those with higher exposure to ACEs were more likely to be food insecure compared to their counterparts. Children with gaps in health insurance over the past 12 months were 1.7 times more likely to be food insecure compared to their peers that had continuous coverage (OR = 1.7; 95% CI: 1.5 – 2.0). Similarly, parental unemployment, receipt of public assistance in family, income less than 200% FPL, parental stress of caregiving, lack of emotional support, and parent’s physical and mental health were all statistically significantly associated with food insecurity. Further, factors such as living in less supportive neighborhood and living in states that did not expand Medicaid were also associated significantly with food insecurity.

**Table 1.**
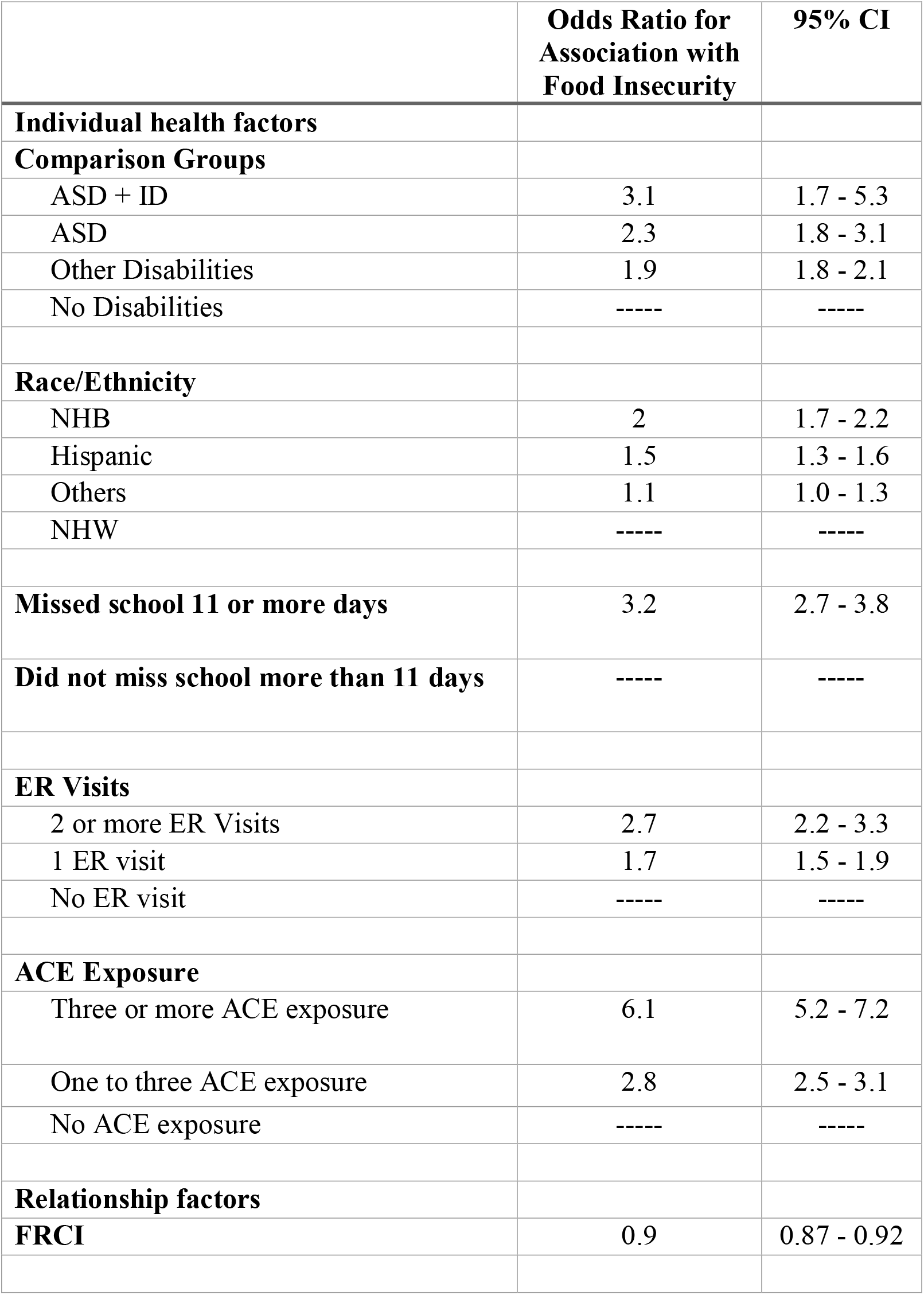
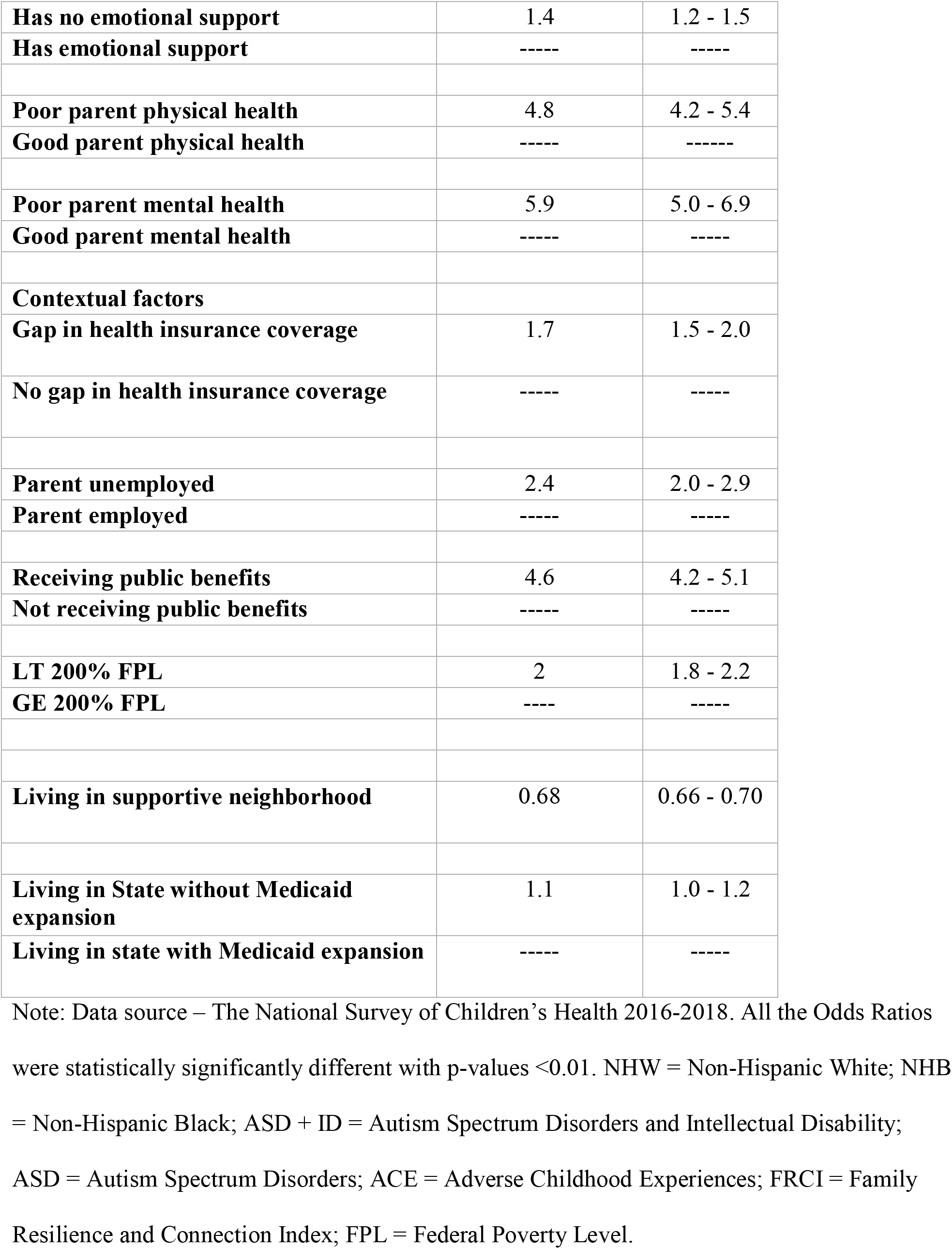
Bivariate analysis of individual health (IH), relationship (R), and contextual (C) factors associated with food insecurity.

### Regression analysis

Table 2 illustrates three logistic regression models explaining food insecurity among children across the comparison groups. The first model includes predictors comprising the IH domain, the second model includes both IH and R domains, and the third model includes factors from the IH, R, and C domains. The model-based adjusted odds ratios, confidence intervals, and percent changes in the Tjur R-squared value are noted in Table 2. In the final model, children with ASD + ID were nearly twice as likely to be food insecure compared to children without disabilities (OR = 1.9; 95% CI: 1.0 – 3.9); children with ASD only were 1.5 times more likely to be food insecure (OR = 1.5; 95% CI: 1.1 – 2.0), and children with other disabilities were 1.3 times more likely to be food insecure (OR = 1.3; 95% CI: 1.2 – 1.5) compared with children without disabilities. The gradient of the severity of food insecurity across the comparison groups was sustained across the different models after adjusting for various factors contained within the IH, R, and C domains where children with ASD + ID were most food insecure, followed by children with ASD, then by children with other disabilities and lastly those with no disability classification. While the race/ethnicity variable was statistically significant in model 1, its significance level reduced in model 2, and its effects were statistically non-significant in model 3. Children that missed school for 11 or more days were nearly two times more likely to be food insecure across all three models. While the strength of association between exposure to adverse childhood experiences reduced with the addition of variables in the R and C domains, its impact remained to substantial in predicting food insecurity. Children of parents that experience poor mental and physical health are 1.5 to 2 times more likely to experience food insecurity compared to their peers with good physical and mental health.

**Table 2.** Multivariate logistic regression models predicting food insecurity across the comparison groups.

| **Independent Variable** | **Model 1**<br>**Individual**<br>**Health factors** | **Model 2**<br>**Individual**<br>**Health &**<br>**Relationship**<br>**factors** | **Model 3**<br>**Individual**<br>**Health,**<br>**Relationship &**<br>**Contextual**<br>**factors** |
| --- | --- | --- | --- |
|  | **Odds Ratio**<br>**(95% CI)** | **Odds Ratio**<br>**(95% CI)** | **Odds Ratio**<br>**(95% CI)** |
| ***Comparison groups*** |  |  |  |
| ASD + ID vs No disabilities | 2.6 (1.6 - 4.3) | 2.1 (1.3 - 3.6) | 1.9 (1.0 - 3.9) |
| ASD vs No disabilities | 2.0 (1.5 - 2.6) | 1.7 (1.3 - 2.2) | 1.5 (1.1 - 2.0) |
| Other disabilities vs No disabilities | 1.4 (1.3 - 1.6) | 1.3 (1.2 - 1.5) | 1.3 (1.2 - 1.5) |
| ***Race/Ethnicity*** |  |  |  |
| NHB vs. NHW | 1.6 (1.4 - 1.9) | 1.5 (1.3 - 1.8) | 1.0 (0.8 - 1.1) |
| Hispanic vs. NHW | 1.5 (1.3 - 1.7) | 1.4 (1.2 - 1.6) | 0.9 (0.8 - 1.1) |
| Others vs. NHW | 1.1 (1.0 - 1.3) | 1.1 (0.9 - 1.2) | 0.9 (0.8 - 1.0) |
| **Missed school > 11 days vs**<br>**Attending school regularly** | 1.9 (1.6 - 2.3) | 1.8 (1.5 - 2.2) | 1.8 (1.5 - 2.2) |
| ***ER Visits*** |  |  |  |
| 2 or more EV vs. None | 1.6 (1.3 - 2.1) | 1.5 (1.2 - 1.9) | 1.1 (0.9 - 1.5) |
| 1 ERV vs. None | 1.4 (1.2 - 1.5) | 1.3 (1.2 - 1.5) | 1.2 (1.0 - 1.3) |
| ***ACE Exposure*** |  |  |  |
| Three or more vs No ACE exposure | 5.0 (4.2 - 5.9) | 3.9 (3.3 - 4.7) | 2.8 (2.3 - 3.3) |
| One to three vs No ACE exposure | 2.5 (2.3 - 2.8) | 2.3 (2.1 - 2.5) | 1.9 (1.7 - 2.2) |
| **FRCI** |  | 0.95 (0.93 - 0.97) | 0.97 (0.95 - 0.99) |
| **No emotional support vs has emotional support** |  | 1.3 (1.1 - 1.4) | 1.1 (1.0 - 1.2) |
| **Poor parental physical health vs good parental physical health** |  | 2.6 (2.2 - 3.1) | 2.0 (1.6 - 2.5) |
| **Poor parental mental health vs good parental mental health** |  | 1.4 (1.1 - 1.8) | 1.4 (1.0 - 1.8) |
| **Gaps HI vs No gaps HI** |  |  | 1.4 (1.1 - 1.7) |
| **Parent unemployed vs employed** |  |  | 1.4 (1.1 - 1.7) |
| **Parental stress index** |  |  | 1.1 (1.0 - 1.1) |
| **Receiving welfare benefits vs. no welfare benefits** |  |  | 3.0 (2.7 - 3.5) |
| **LE 200 % FPL vs GT 200% FPL** |  |  | 1.2 (1.1 - 1.4) |
| **Supportive neighborhood** |  |  | 0.76 (0.73 - 0.79) |
| **No Medicaid Expansion vs Medicaid Expansion** |  |  | 1.1 (1.0 - 1.2) |
| *C- statistic* | *0.707* | *0.735* | *0.784* |
| *Tjur Rsquare* | *0.0925* | *0.1288* | *0.183* |
| *% Change in r-square* |  | *28.2%* | *29.6%* |

Note: Data source – The National Survey of Children’s Health 2016-2018. All the Odds Ratios were statistically significantly different with p-values <0.01 except for race/ethnicity. NHW = Non-Hispanic White; NHB = Non-Hispanic Black; ASD + ID = Autism Spectrum Disorders and Intellectual Disability; ASD = Autism Spectrum Disorders; ACE = Adverse Childhood Experiences; FRCI = Family Resilience and Connection Index; FPL = Federal Poverty Level.

Children from families that received public assistance (i.e., SNAP, reduced price breakfast or lunch, or other cash assistance) were nearly three times more likely to be food insecure compared to their peers that did not receive such benefits. Living in a supportive neighborhood was a protective factor in reducing the likelihood of food insecurity by 24% compared with families that lived in a non-supportive neighborhood. Children living in states that did not expand Medicaid were 110% more likely to experience food insecurity.

Examining Tjur R-squared values, it is evident that the addition of variables from the R domain improved the prediction of food insecurity by 28% in the model with IH factors. The addition of variables from the C domain further improved the prediction by another one third (or 30%) in the model consisting of both IH and R domain variables. The c-statistic also improved across the three models (0.707, 0.735, 0.784).

## Discussion

Based on three years of nationally representative data, the prevalence of food insecurity was highest in households of children with ASD + ID, followed by those with children with ASD, and then by those with other disabilities. Households of children with no disabilities were least likely to experience food insecurity. Specifically, approximately four in every ten families of children with ASD + ID or ASD only are food insecure, based on these data. Initiatives that address this problem in the households of children with ASD are urgently needed.

Food insecurity has remained to be a major challenge in the U.S. since the onset of the Great Recession of 2009, and the disparities in its prevalence between individuals with and without disabilities have remained high. The current COVID19 pandemic is likely to worsen food security, as food insecurity has been strongly correlated with unemployment, and the pandemic has already led to higher unemployment (Gundersen & Ziliak, 2018; Kinsey, Kinsey, & Rundle, 2020). A recent study illustrated a three-fold increase in rates of food insecurity in U.S. households with children since the beginning of this public health emergency (Schanzenbach & Pitts, 2020). Qualitatively, many families are not able to buy enough food items from stores and have reported the need to travel to multiple stores. Challenges in obtaining food from food pantries or stores that accept food stamps have also increased during the pandemic (Kinsey, Oberle, Dupuis, Cannuscio, & Hillier, 2019). It is evident that these challenges compound the additional demands of caregiving that parents experience from disruption in access to usual source of care during the pandemic (Colizzi et al., 2020; Jeste et al., 2020).

Interestingly, while various individual health, relationship, and contextual factors were associated with the likelihood of food insecurity in this study, some factors showed stronger and more consistent associations across models. For example, exposure to three or more ACEs was positively correlated with the likelihood of food insecurity after multivariate adjustments across all the models. Alternatively, the statistical significance of the race/ethnicity variable was substantially reduced as additional factors were added into the model. One of the important reasons is that the race/ethnicity variable is highly correlated to variables in the R and C domains indicative of socio-economic vulnerabilities. For example, families of racial/ethnic minorities have higher participation rates in public assistance programs as explained by factors such as earnings, family structure, education, and other variables, representing more disadvantaged status (Moffitt & Gottschalk, 2001). Also, structural racism manifesting through lack of equitable employment opportunities and a higher probability of discrimination based on race/ethnic background (related to the R domain of the sociotype) contribute to a larger proportion of racial/ethnic minorities relying on public assistance and food insecurity (Burke et al., 2018; Odoms-Young & Bruce, 2018).

Exposure to adverse childhood experiences (ACE) was related to higher odds of food insecurity. In addition to the lasting detrimental impact of ACE on physical and mental health well-being across children’s lifespan, longitudinal studies have indicated that childhood exposure to ACE predicts a three- or four-fold increase in food insecurity among young adults (Testa & Jackson, 2020). It is likely that the COVID19 pandemic has increased such exposure and potentially the intensity of its impact on the psychosocial well-being of children with ASD (Bryant, Oo, & Damian, 2020; Gruber et al., 2020). Cumulative stress from the impact of the pandemic on caregivers and the additional burden of caregiving caused by the closure of schools and other sources of child care is likely to increase the risk of ACE exposures (Brown, Doom, Lechuga-Peña, Watamura, & Koppels, 2020). These trends especially disadvantage children with ASD who were three times more likely to experience ACEs prior to the pandemic. (Fegert, Vitiello, Plener, & Clemens, 2020; Kerns & Lee, 2015). Interventions targeting caregivers supports are needed, especially for families of children with ASD, to equip them with strategies and skills for positively engaging children, and improve their access to psychosocial resources to prevent or mitigate food insecurity and other basing needs such as housing (Eshraghi et al., 2020; Scahill et al., 2016). This research also underscores a need to ensure health care providers, school personnel, and social workers are equipped to screen for ACEs, and implement mitigating strategies to reduce their impact by adopting practices rooted in positive psychology and trauma-informed care (Ranjbar & Erb, 2019; Sciaraffa, Zeanah, & Zeanah, 2017).

After controlling for other factors, the receipt of public assistance continued to disadvantage families, increasing their likelihood of becoming food insecure. While it is likely that most financially vulnerable families qualify for public assistance, the threefold higher rates of food insecurity among families receiving public assistance raise questions about the effectiveness of these programs in reducing food insecurity in vulnerable families of children with ASD. Notably, SNAP, implemented nationally since 1974, continues to play an important role in supporting access to food for low-income families. However, the program is designed to supplement only 2/3rd of the monthly food costs for eligible households, leaving many families on SNAP food insecure (Gregory & Smith, 2019; Kinsey et al., 2019). Estimates indicate that more than 50% of families receiving SNAP benefits remain food insecure, and many eligible families do not apply for SNAP benefits due to restrictive eligibility and recertification requirements, relatively low benefit levels for those closer to the upper limits of income eligibility, and social stigma of receiving public benefits (Wu & Eamon, 2010). However, prior econometric studies illustrated that the receipt of SNAP benefits reduces the likelihood of food insecurity (Ratcliffe & McKernan, 2010). In the wake of the COVID19 pandemic, Congress has passed the Family First Coronavirus Response Act (FFCRA), which allows states to increase SNAP benefits to the maximum allowable thresholds for recipients. However, it is likely that the pandemic may have significantly impeded SNAP recipient’s ability to strategically shop and stockpile needed food. Store closures, restrictions in shopping hours, restrictive state policies preventing online food purchase, and disruption in food supply chains, in general, are likely to impact access to needed food for these vulnerable families during the pandemic. Online shopping for food continues to be limited for families receiving SNAP benefits despite easing restrictions since May 2020, and pose additional challenges experienced by the families. Investments in leveraging technology to support low-income families to shop efficiently are urgently needed to prevent food insecurity among households of children with ASD. While a handful of the states have explored additional flexibility in administering SNAP benefits and leveraged Medicaid waivers (e.g., section 1115, 1135, or 1915c) to provide access to home-delivered meals, the impact of these programs will provide further direction for future program and policy innovations. Prior research found that expanding SNAP benefits reduced monthly inpatient costs by about 73%, and similarly reduced rates of inpatient admissions for Medicaid enrollees (Sonik, 2016). These data point to an optimistic picture of the positive impact of SNAP expansion efforts on health care systems and may further benefit families of children with ASD. Given the cost-benefit of food assistance programs returning savings in health care expenditures, health care payors, both public and private, should explore options to improve access to adequate nutrition for families of individuals with ASD.

The breakdown of community supports resulting from measures to ensure social distancing, and quarantining, while necessary to contain COVID19 spread, may disrupt the social fabric of support for families of individuals with ASD. In the current analysis, living in supportive neighborhoods reduced the likelihood of food insecurity by 21%-27%. Prior studies indicate that social cohesion reduces the risk current and future risk of food insecurity among SNAP beneficiaries (J. Denney, Kimbro, Heck, & Cubbin, 2016). Since the families of children with ASD already report a high-level of social isolation (Lai, Goh, Oei, & Sung, 2015), having a network of supports become crucial and consequential for families during the COVID19 pandemic. Additionally, the analysis indicated increased vulnerabilities in families where parents lacked source of emotional support. Efforts at expanding programs that build social networks for families are needed to mitigating challenges and promoting food insecurity (Catalano, Holloway, & Mpofu, 2018).

Living in states that expanded Medicaid under the ACA was a protective factor in food insecurity. Hilmmelstein (2019), using the Current Population Survey data, demonstrated that expansion of access to Medicaid under the Affordable Care Act reduced very low food insecurity among individuals in Medicaid-expansion states by about 13% compared with an increase in very low food insecurity among individuals with non-expansion states (G. Himmelstein, 2019). It is also likely that the states that expanded Medicaid access by 2016 were also likely to be the states that had, to a varying degree (Velott et al., 2016), implemented state-wide HCBS waivers funding services for individuals with ASD. Future research should consider examining the characteristics of HCBS waivers and it’s relevance in the context of addressing food insecurity among families of children with ASD in states that did not choose to expand Medicaid under the ACA. The COVID19 relief efforts should provide supports to programs such as autism-specific HCBS waivers to advance access to needed services, in addition to encouraging states to expand Medicaid access.

### Limitations

The current analysis is limited by the survey-based cross-sectional data collection approach used in the NSCH. This approach is not able to rule in/out causative factors, nor can it clearly delineate the temporal pattern of how factors at each level of analysis lead to food insecurity. The reported association between the individual health, relationships, and contextual factors impacting food insecurity are at best correlational. While the ‘sociotype’ framework provided a useful way to examine food insecurity and its contributing factors, the process in which different sociotypes manifest is better understood with approaches such as the structural equations models and latent class analysis accompanied with longitudinal data collection (Jock et al., 2020; Mesoudi et al., 2017). Additionally, multi-level modeling strategy can help identify how macro-level factors such as specific policies (e.g., Medicaid expansion, ASD-specific Medicaid waivers, etc.) impact food security at the family-levels.

The ascertainment of the comparison groups – ASD + ID, ASD, Other Disability, and No Disability, was based on the parent-reported information on diagnosis for their children. Prior studies have demonstrated that the prevalence of ASD based on the NSCH has been higher compared to prevalence estimates reported through the clinical and education records-based surveillance approach used by the Centers for Disease Control and Prevention (CDC) (Kogan et al., 2018). However, parent-reported ASD diagnoses have been found to be a valid approximation to clinical ASD diagnoses in survey contexts, and unreliability in group classifications is likely to attenuate relationships between these groups and food security, making this a conservative analysis (Daniels et al., 2012).

On the one hand, the strategy of utilizing food insecurity as a binary variable without stratifying this into additional categories (e.g., low food security, very low food security, etc.) is consistent with a lack of differentiation in prior studies on the other it could reduce predictive validity and induce heterogeneity in the analyses. However, studies have indicated that households with marginal food insecurity are similar to the food-insecure households and have similar levels of negative impact on health outcomes for children and their caregivers (Cook et al., 2013). Also, prior research has indicated equivalence for a simpler two-item measure for identifying food insecurity in comparison with the 18-item measure providing additional support for the strategy utilized in this analysis (Hager et al., 2010).

### Implications

Food insecurity is a significant vulnerability among families of children with ASD + ID and those with ASD, and efforts to improve the wellbeing of individuals with ASD must therefore address food insecurity. Besides the COVID19 pandemic, having disproportionately impacted individuals with ASD and other developmental disorders (Colizzi et al., 2020; Lima, Barros, & Aragao, 2020) is likely to engender food insecurity creating negative feedbacks that snare their social inclusion and development. The sociotype framework provides an opportunity to broaden the challenge of food insecurity as an economic issue to a more bio sociopsychological challenge that can be remedied effectively through a multi-level approach rooted in public health principles.

## Data Availability

Data for the manuscript are publicly available from the U.S. Census Bureau.

https://www.census.gov/programs-surveys/nsch/data.html

